# Preoperative Systemic Immune-Inflammation Index and Clinical Outcomes After Coronary Artery Bypass Grafting: A Systematic Review and Meta-analysis

**DOI:** 10.64898/2026.10.17.26365801

**Authors:** Saeed Nourian, Kimia Jalalian Rad, Mina Ghasemi, Mohadese Mosavi Mirkalaie, Nastaran Tayebi, Mohammad Moein Ashrafi, Mohammad Hossein Nourallahzadeh, Ali Reza Norouzi

## Abstract

**Background:** The systemic immune-inflammation index (SII) is a routinely available blood-derived marker with potential prognostic relevance after coronary artery bypass grafting (CABG). We evaluated associations between preoperative SII and postoperative outcomes.

**Methods:** PubMed, Embase, Scopus and Web of Science were searched to 1 October 2026, with supplementary searches through 14 October 2026. Peer-reviewed observational studies evaluating preoperative SII in adults undergoing CABG were eligible. Methodological quality was assessed using the Newcastle–Ottawa Scale. Compatible categorical associations and exploratory adjusted continuous mortality effects were pooled separately using restricted maximum-likelihood random-effects models; other findings were summarized descriptively.

**Results:** Thirty-one reports were included, with an aggregate sample of 30,976 and possible cohort overlap; five contributed to meta-analysis. Two off-pump cohorts comprising 1,378 patients and 180 atrial-fibrillation events yielded an imprecise crude association with postoperative atrial fibrillation (OR 1.60, 95%CI 0.53–4.82; I^2^=68%). An exploratory crude synthesis indicated greater intra-aortic balloon pump use with higher SII (OR 2.45, 95%CI 1.16–5.18; I^2^=51%). Two cohorts with differing follow-up yielded an exploratory adjusted mortality HR of 1.11 (95% Wald CI 1.05–1.18) per study-specific SD increase in log-SII. In one 624-patient cohort, high SII was associated with postoperative acute kidney injury after adjustment (OR 3.35, 95% CI 2.25–4.97).Apparent atrial-fibrillation discrimination varied (AUC 0.587–0.946).

**Conclusions:** Preoperative SII showed associations with selected adverse outcomes after CABG, supporting further evaluation as an accessible prognostic marker. Its incremental value beyond established clinical risk models requires validation.

## 1. Background

Coronary artery bypass grafting (CABG) remains an important revascularization strategy for selected patients with complex coronary artery disease [1, 2]. Preoperative risk assessment informs treatment decisions and perioperative planning. Established models, including the Society of Thoracic Surgeons risk models and the European System for Cardiac Operative Risk Evaluation II (EuroSCORE II), incorporate clinical and procedural factors to estimate operative risk [3, 4]. Inflammatory biomarkers have also been associated with morbidity and mortality after cardiac surgery [5]. However, uncertainty about the causal relationship between immune activation and organ injury reinforces the need to distinguish prognostic associations from mechanisms of disease [6].

The systemic immune-inflammation index (SII), calculated as platelet count multiplied by neutrophil count and divided by lymphocyte count, combines three routinely measured blood-cell populations [7, 8]. SII has subsequently been associated with cardiovascular outcomes in patients with coronary artery disease undergoing percutaneous coronary intervention [9]. These findings support its investigation in surgical populations, although its prognostic role after CABG requires direct evaluation. Because it uses routine laboratory measurements, SII offers a practical candidate for evaluation alongside established clinical risk factors.

CABG studies have examined preoperative SII in relation to composite postoperative complications [10], atrial fibrillation [11–13], acute kidney injury [14], and prolonged mechanical ventilation and intensive care unit stay [15, 16]. Investigations have also addressed postoperative delirium in patients with cerebral infarction and longer-term cardiovascular outcomes in selected CABG populations [17, 18]. More recent evidence includes a large isolated-CABG cohort evaluating blood count-derived indices in relation to perioperative and longer-term outcomes [19]. An inverse association between preoperative SII and wound infection was reported in diabetic patients, indicating that the direction of association may vary across populations and endpoints [20].

An earlier meta-analysis evaluated SII for postoperative atrial fibrillation following cardiac surgery, including CABG and other procedures [21]. Its endpoint-specific scope does not address the broader range of CABG outcomes. Differences in sampling intervals, thresholds, outcome definitions, follow-up, and clinical adjustment complicate interpretation. Furthermore, prognostic association alone does not establish accurate prediction or incremental value beyond established risk models. Postoperative measurements may reflect surgical exposures or evolving complications and therefore address a different prognostic question. Focusing on measurements obtained before surgery provides a defined assessment of baseline prognosis. We therefore aim to systematically review the association between preoperative SII and clinical outcomes after CABG in adults, particularly mortality, cardiovascular events, and major postoperative complications.

## 2. Methods

### 2.1 Review design and reporting

This systematic review evaluated preoperative systemic immune-inflammation index (SII) as a prognostic factor for clinical outcomes after coronary artery bypass grafting (CABG), following methodological guidance for reviews of prognostic factor studies [22]. Reporting followed the Preferred Reporting Items for Systematic Reviews and Meta-Analyses (PRISMA) 2020 statement and PRISMA-S [23, 24]. The PRISMA checklist is provided in Supplementary Table S1. The review protocol was registered in PROSPERO (registration number: CRD420261533473) and is available at https://www.crd.york.ac.uk/PROSPERO/view/CRD420261533473.

### 2.2 Eligibility criteria

Eligible studies included adults aged 18 years or older undergoing CABG and evaluated SII measured before surgery in relation to at least one subsequent clinical outcome. Both on-pump and off-pump CABG and elective and urgent procedures were eligible. Studies of mixed cardiac surgical populations or combined procedures were eligible only when data for isolated CABG were separately extractable. Studies restricted to particular CABG populations, such as patients with diabetes or previous cerebral infarction, were eligible, and these restrictions were recorded. Eligible designs comprised prospective or retrospective cohort studies and case-control or nested case-control studies in which SII measurement preceded the outcome. Secondary analyses of trial or registry data were eligible if they provided an extractable assessment of preoperative SII and subsequent CABG outcomes. SII could be assessed continuously or categorically using study-defined thresholds or exposure groups. Eligibility did not depend on statistical significance, the direction of association or the availability of an adjusted estimate.

Studies evaluating only intraoperative or postoperative SII, or changes in SII without a separately extractable preoperative association, were excluded. Studies reporting both preoperative and later measurements were eligible, but extraction focused on the preoperative exposure. Studies of nonsurgical coronary disease, non-CABG surgery without separable CABG data and measurements obtained only after the outcome were excluded. Only original studies published as full-length articles in peer-reviewed journals were eligible. Conference abstracts, theses, dissertations, preprints and unpublished reports were excluded. No minimum sample size or language restriction was imposed.

### 2.3 Definition of the prognostic factor

The exposure of interest was preoperative SII, calculated as platelet count × neutrophil count / lymphocyte count [7]. Eligible blood samples were obtained before CABG began. The reported sampling interval and its timing relative to hospital admission, anesthesia and surgery were recorded whenever available. When several preoperative samples were reported, the investigators’ designated baseline value was identified, and other eligible preoperative time points were documented. Blood-count and SII units, transformations or standardization, categorical boundaries, reference categories and cutoff-selection methods were extracted. Studies reporting SII alongside other inflammatory indices were eligible only when SII-specific information was available.

### 2.4 Clinical outcomes and follow-up

The primary outcome domain was all-cause mortality after CABG, with in-hospital, 30-day and longer-term endpoints considered separately. Secondary domains included cardiovascular mortality; major adverse cardiovascular or cerebrovascular events; postoperative atrial fibrillation; acute kidney injury or renal replacement therapy; myocardial infarction; stroke; postoperative delirium; pulmonary complications; wound infection, other infection or sepsis; graft failure or impaired graft patency; prolonged mechanical ventilation; and intensive care unit or hospital length of stay. Perioperative intra-aortic balloon pump use was examined as an exploratory support endpoint, with timing and indications recorded when reported. Other clinically defined postoperative complications were recorded without replacing the primary outcome.

Original study definitions, ascertainment methods and follow-up intervals were retained. Definitions of atrial fibrillation, kidney injury, infection and prolonged ventilation were extracted rather than assumed to be equivalent across studies. Composite endpoint components were documented. Outcome timing was referenced to CABG whenever possible, and mortality within 30 days was distinguished from mortality during longer follow-up. Biomarker changes, operative duration and cardiopulmonary bypass duration alone did not qualify as postoperative clinical outcomes.

### 2.5 Information sources and search strategy

PubMed, Embase, Scopus and the Web of Science Core Collection were searched from inception to 1 October 2026. Searches combined free-text terms for SII with terms for CABG and surgical coronary revascularization, supplemented by controlled vocabulary where available. Outcome terms and study-design filters were not required. No publication-date or language restrictions were applied. Database-specific search strategies are provided in Supplementary Table S2. Reference lists of eligible reports and relevant systematic reviews were manually screened to identify additional potentially eligible studies.

### 2.6 Study selection and linked reports

Search records were exported to EndNote 21, and duplicates were removed using automated matching followed by manual verification. Two reviewers, M.G. and K.J.R., independently screened titles and abstracts and assessed potentially eligible full texts. Before formal screening, the reviewers piloted the eligibility criteria on a sample of records. Disagreements were resolved through consultation with a third reviewer, M.M.M. Reports were compared for possible cohort overlap using author groups, centers, recruitment dates, sample sizes, participant characteristics and data-source identifiers. Multiple publications from the same cohort were linked under a common study identifier where overlap could be established. Reports providing distinct outcomes or additional follow-up were retained as sources for that study. Report counts were distinguished from independent study counts, and unresolved overlap was documented. Investigators were contacted when uncertainty about recruitment periods, CABG subgroups or cohort overlap prevented a reliable decision.

### 2.7 Data extraction and verification

Two reviewers, N.T. and M.H.N., independently extracted data using a standardized form. Extracted data were cross-checked against the original sources, and disagreements were resolved through discussion. Disagreements that remained unresolved were adjudicated by a third reviewer, S.N. Extracted characteristics included publication details, country, center and data source, study design, recruitment dates, eligibility criteria, sample size, age, sex, relevant comorbidities, operative urgency, surgical approach and concomitant procedures. SII measurement timing, calculation, units, exposure categories and cutoff derivation were recorded alongside outcome definitions, ascertainment, event counts, follow-up and completeness of outcome data.

For each eligible SII–outcome association, reviewers recorded the effect measure and estimate, confidence interval, exposure scale or comparison, reference group, model population and adjustment variables. Adjusted and unadjusted estimates were recorded separately when available. Discrimination measures, including the area under the receiver operating characteristic curve, and information on calibration, validation or incremental value beyond clinical models were extracted separately from association estimates. Continuous outcome summaries and corresponding group sizes were also recorded. Data were obtained from the main text, tables, figures and available supplementary materials. Source locations were recorded for extracted values. Discrepancies between abstracts and main reports, numerical inconsistencies and uncertain model labels were checked against the original sources and reviewed by the third reviewer when necessary. Source inconsistencies that could not be resolved were explicitly documented, without correction based on unsupported assumptions. Missing information was marked as not reported.

### 2.8 Methodological quality and risk of bias assessment

Methodological quality was assessed independently by two reviewers, M.G. and M.M.M., using the Newcastle–Ottawa Scale (NOS), applying the cohort or case-control version as appropriate [25]. The NOS assesses selection, comparability and outcome ascertainment for cohort studies, or exposure ascertainment for case-control studies. Selection contributes up to four stars, comparability up to two stars and outcome or exposure ascertainment up to three stars, giving a maximum total of nine. Domain and total scores were recorded for the index outcome specified for each report. Disagreements were resolved through discussion by a third reviewer, N.T. Total scores were not interpreted as categorical risk-of-bias judgments. Assessments are provided in Supplementary Table S6.

### 2.9 Statistical analysis

Categorical associations with sufficiently comparable exposure contrasts, outcome definitions and follow-up intervals were synthesized as odds ratios (ORs) with 95% confidence intervals (CIs). Exploratory categorical analyses incorporating estimates with unspecified endpoint-specific adjustment were distinguished from crude-only syntheses. Separately, adjusted all-cause mortality hazard ratios (HRs) expressed per study-specific standard-deviation increase in log-transformed SII were combined in an exploratory synthesis, retaining differences in follow-up horizons and covariate adjustment. Standard errors for reported ORs and HRs were approximated as [ln(upper confidence limit) − ln(lower confidence limit)]/(2×1.96). Estimates were pooled on the logarithmic scale using inverse-variance random-effects models, with between-report variance estimated by restricted maximum likelihood (REML). Two-report analyses and analyses with zero estimated between-report variance used Wald CIs; three-report analyses with positive estimated between-report variance used Hartung–Knapp CIs. Heterogeneity was summarized using τ^2^ and I^2^, with I^2^ calculated from the estimated between-report variance and typical within-report sampling variance. Continuous and categorical contrasts and ORs and HRs were synthesized separately. Explicitly adjusted effects were analyzed separately from crude effects. Exploratory categorical analyses additionally combined crude estimates with source-reported estimates whose endpoint-specific adjustment was unspecified. Sensitivity analyses examined common-effect models, Paule–Mandel variance estimation, alternative CI methods, leave-one-out omission and crude risk ratios, as applicable. The adjusted mortality synthesis additionally used a modified Hartung–Knapp interval with a variance floor of 1 and k−1 degrees of freedom. Other associations and discrimination measures were summarized descriptively.

## 3. Results

### 3.1 Study selection

The database searches identified 777 records: 102 from PubMed, 197 from Embase, 301 from Scopus, and 177 from Web of Science. After removing 232 duplicates, 545 records underwent title and abstract screening, and 491 were excluded. All 54 reports sought for full-text assessment were retrieved. Of these, 23 were excluded because of an ineligible population or procedure, including nonextractable CABG-specific data (n=7); absence of a separately extractable preoperative SII assessment (n=3); absence of an eligible postoperative clinical outcome (n=4); or an ineligible study design or publication type (n=9). Ultimately, 31 reports were included in the systematic review. Five contributed to at least one quantitative synthesis: three to categorical analyses and two additional reports to the exploratory adjusted continuous mortality analysis. Figure 1 presents the updated study-selection process.

**Figure 1.**
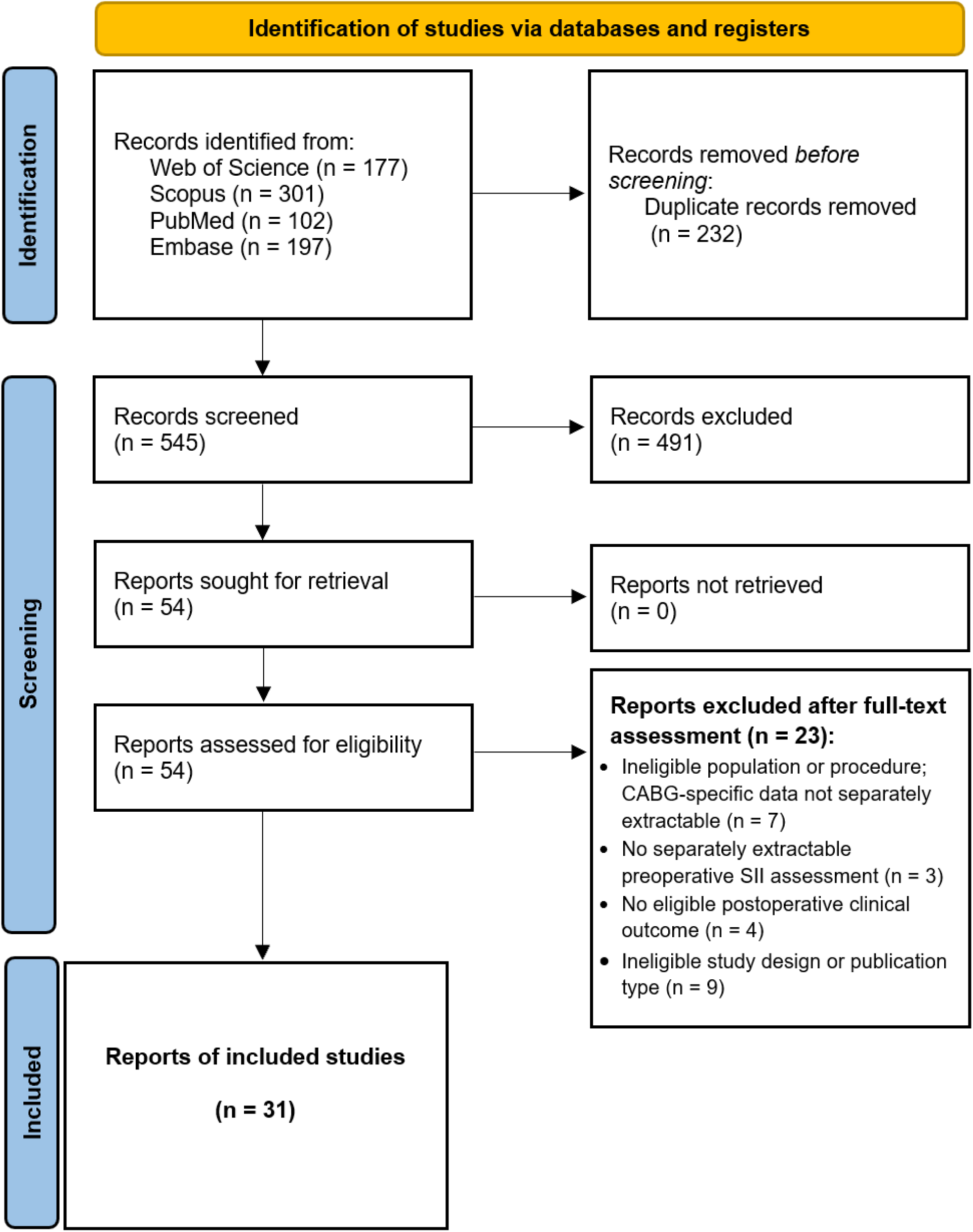
PRISMA 2020 flow diagram of the study selection process.

### 3.2 Characteristics of the included reports

Thirty-one reports evaluated preoperative SII in relation to clinical outcomes after CABG [10–20, 26–45], with an aggregate sample size of 30,976. Reports were published between 2021 and 2026 and originated from Türkiye (19 reports), China (four), Poland (three), Indonesia (two), India (two) and the United States (one). Most used retrospective cohort analyses, and one used a mortality-enriched nested case–control sample. Report-level sample sizes ranged from 50 to 9,046 (Table 1). Populations included selected elective cohorts and patients restricted by diabetes, ventricular function, previous cerebral infarction, coronary anatomy or younger age. The Polish Diagnostics and Cells reports probably describe overlapping populations [34, 35]. Potential overlap also requires consideration between the Indian Dey and Magoon reports and between the Chinese Osmanaj and Xue reports. SII was derived from preoperative blood counts, although sampling intervals and reporting completeness varied.

**Table 1.** Characteristics of the included reports.

| Report | Country | n | Design | Pump | Population | Follow-up |
| --- | --- | --- | --- | --- | --- | --- |
| Dey et al. 2021 [10] | India | 1007 | Retrospective cohort | Off-pump | Elective off-pump CABG | 30 days |
| Selcuk et al. 2021 [11] | Türkiye | 391 | Retrospective cohort | On-pump | Isolated on-pump CABG | Postoperative period; exact surveillance window NR |
| Ata & Abanoz 2021 [28] | Türkiye | 283 | Retrospective cohort | On-pump | Isolated CABG with right coronary artery disease | In-hospital |
| Uguz et al. 2022 [13] | Türkiye | 116 | Retrospective cohort | On-pump | Elective isolated on-pump CABG | Until hospital discharge |
| Topal et al. 2022 [12] | Türkiye | 722 | Retrospective cohort | Off-pump | Elective isolated off-pump CABG | Until discharge |
| Urbanowicz et al. 2022 [36] | Poland | 171 | Retrospective cohort | Off-pump | Chronic coronary syndrome; isolated off-pump revascularization | 8 years |
| Yilmaz et al. 2023 [38] | Türkiye | 392 | Retrospective cohort | On-pump | On-pump CABG | Before hospital discharge |
| Erol & Civelek 2025 [29] | Türkiye | 100 | Retrospective cohort | On-pump | Isolated on-pump CABG with EF $\leq 40\%$ (Methods); abstract $< 40\%$ | Postoperative admission; precise window NR |
| Aykut et al. 2023 [14] | Türkiye | 624 | Retrospective cohort | On-pump | Isolated on-pump CABG | AKI within 7 days; other outcomes including 30-day mortality |
| Alagha et al. 2023 [27] | Türkiye | 99 | Retrospective cohort | Mixed | Isolated elective CABG; 69 on-pump and 30 off-pump | 30 days |
| Önder et al. 2026 [31] | Türkiye | 500 | Retrospective cohort | On-pump | Adult isolated elective on-pump CABG | In-hospital / early postoperative; fixed day limit NR |
| Öntaş and Gözüaçık Rüzgar 2026 [20] | Türkiye | 300 | Retrospective cohort | On-pump | Diabetic adults undergoing isolated elective on-pump CABG | Index admission or $\leq 30$ days after discharge (Methods); abstract states 30 days after surgery |
| Jacquemyn et al. 2026 [19] | USA | 9046 | Retrospective cohort | Mixed | Adults undergoing isolated CABG; elective and non-elective | Early: 30 days/index admission; median long-term 5.3 years [IQR 2.4–9.2] |
| Parmana et al. 2023 [15] | Indonesia | 371 | Retrospective cohort | Off-pump | Adults undergoing non-emergency off-pump CABG | Mortality: 30 days; other outcomes during perioperative admission |
| Sanrı and Aslan 2026 [32] | Türkiye | 120 | Retrospective cohort | On-pump | Elective isolated CABG with CPB | Postoperative hospital stay; no long-term outcome analysis |
| Parmana et al. 2024 [16] | Indonesia | 1056 | Retrospective cohort | On-pump | Non-emergency on-pump CABG; valve surgery excluded | Mortality at 30 postoperative days; other outcomes perioperative admission |
| Urbanowicz et al. 2022 [34] | Poland | 175 | Retrospective cohort | Off-pump | Diabetic subgroup after elective isolated off-pump CABG | $3.7 \pm 1.5$ years; mean/median label inconsistent |
| Zhao et al. 2024 [18] | China | 321 | Retrospective cohort | Off-pump | OPCAB with neurologist-confirmed pre-existing cerebral infarction | 5 postoperative days |
| Toprak et al. 2026 [33] | Türkiye | 245 | Nested case–control | On-pump | Elective isolated on-pump CABG; all 51 deaths and 194 randomly sampled surviving controls | Index hospitalization |
| Wang et al. 2024 [17] | China | 294 | Retrospective cohort | Not reported | New-onset ischemic cardiomyopathy with symptomatic HFpEF undergoing CABG | Median 34.98 months |
| Urbanowicz et al. 2022 [35] | Poland | 538 | Retrospective cohort | Off-pump | Consecutive OPCAB for complex chronic coronary syndrome | Mean $4.7 \pm 1.7$ years |
| Vezir and Ertürk Tekin 2026 [37] | Türkiye | 1034 | Retrospective cohort | On-pump | Adults undergoing elective isolated on-pump CABG; complete data; postoperative stay $\geq 48$ h | Postoperative hospital stay |
| Göç and Bağış 2026 [30] | Türkiye | 50 | Retrospective cohort | On-pump | Elective isolated on-pump CABG; adults aged 20–85 years | AKI: first 48 hours; other early postoperative outcomes |
| Abanoz and Engin 2021 [26] | Türkiye | 297 | Retrospective cohort | On-pump | Elective isolated CABG | 1 month |
| Magoon et al. 2023 [39] | India | 899 | Retrospective cohort | Off-pump | Elective OPCABG; patients with pre-existing arrhythmia and concomitant valve surgery excluded | AF during the first postoperative week |
| Dolapoglu and Avcı 2024 [40] | Türkiye | 527 | Retrospective cohort | On-pump | Elective CABG for stable coronary artery disease; concomitant valve surgery excluded | Median 4.6 years; conflicting 12-month statement in Methods |
| Atasoy and Guven 2025 [41] | Türkiye | 223 | Retrospective cohort | Off-pump | Elective first-time isolated CABG; age >18 to ≤50 years | Rhythm monitoring through discharge |
| Osmanaj et al. 2026 [42] | China | 3,481 | Retrospective cohort | NR | Isolated CABG without preoperative AF; emergency and concomitant cardiac procedures excluded | AF within seven postoperative days |
| Şahin and Yeniay 2026 [44] | Türkiye | 618 | Retrospective cohort | NR | Adult isolated CABG; emergency and combined cardiac procedures excluded | In-hospital and one-year mortality |
| Xue et al. 2026 [43] | China | 6,549 | Retrospective multicenter cohort | Off-pump | Adults undergoing elective OPCABG at eight centers | In-hospital postoperative cerebral infarction |
| Yüksel et al. 2024 [45] | Türkiye | 427 | Retrospective cohort | Off-pump | Isolated off-pump CABG, October 2021–December 2023; pre-existing AF, emergency/redo surgery and concomitant procedures excluded | AF surveillance through hospital discharge |

n is the report-level analyzed sample, not always the SII-specific model denominator. Jacquemyn: SII available in 8,993/9,046. Wang: SII model developed in 176 of 294 patients. Urbanowicz Diagnostics: 175-patient diabetic analysis rather than the entire source cohort. Toprak: 245 sampled participants from 874 eligible patients. NR, not reported.

### 3.3 All-cause mortality

Categorical associations with 30-day mortality and adjusted continuous associations with mortality over longer follow-up were synthesized separately. The exploratory categorical synthesis combined a calculated crude OR with a source-reported OR whose endpoint-specific adjustment was unspecified. The separate exploratory synthesis of adjusted HRs combined associations per study-specific SD increase in log-transformed SII across differing follow-up horizons (Figure 2C; Tables 2 and 3; Supplementary Tables S3 and S7).

**Figure 2.**
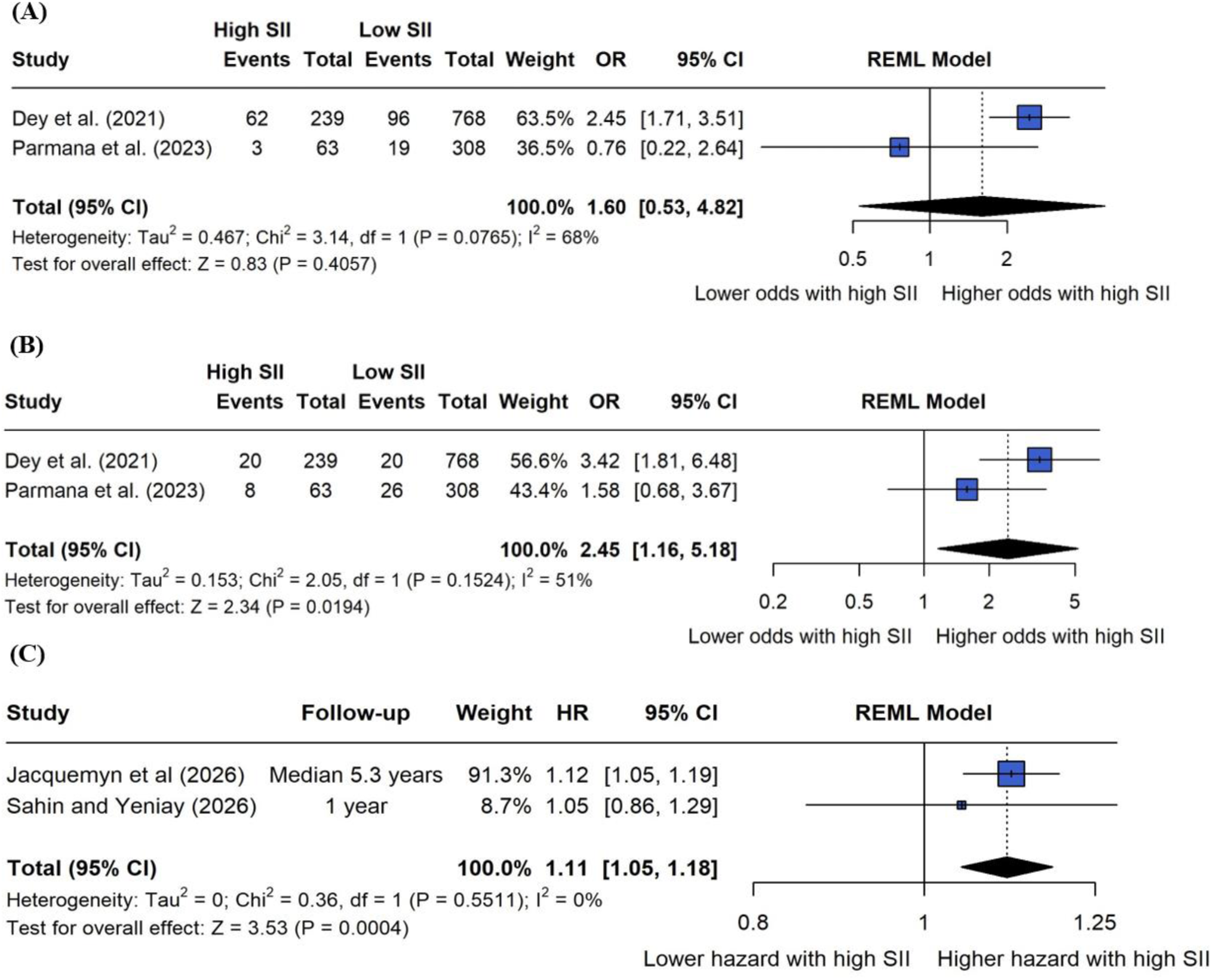
Associations between preoperative systemic immune-inflammation index and clinical outcomes after coronary artery bypass grafting. (A) Crude categorical associations with postoperative atrial fibrillation. (B) Exploratory crude categorical associations with perioperative intra-aortic balloon pump use. Panels A and B compare SII ≥878.057 with SII <878.057 in two off-pump cohorts. (C) Exploratory adjusted associations with all-cause mortality per study-specific standard-deviation increase in log-transformed SII. All panels use inverse-variance random-effects models with restricted maximum-likelihood estimation and Wald 95% confidence intervals. Square areas reflect study weights, horizontal lines indicate study-specific confidence intervals, and diamonds represent pooled estimates and their confidence intervals. Dashed vertical lines indicate the null value of 1. Each panel represents a separate meta-analysis. Panel C combines different follow-up horizons and covariate adjustment sets. CI, confidence interval; HR, hazard ratio; OR, odds ratio; SII, systemic immune-inflammation index.

**Table 2.** Selected reported adjusted SII associations.

| Report | Outcome | Effect and contrast | Reported estimate (95% CI) | Interpretation or restriction |
| --- | --- | --- | --- | --- |
| Jacquemyn et al. 2026 [19] | Long-term mortality | HR per SD of log-SII | 1.12 (1.05–1.19) | included in the exploratory adjusted continuous mortality synthesis; median follow-up 5.3 years. |
| Jacquemyn et al. 2026 [19] | Long-term MACCE | HR per SD of log-SII | 1.08 (1.03–1.13) | Adjusted; distinct from other MACE definitions |
| Aykut et al. 2023 [14] | AKI within seven days | OR; >651.37 vs ≤651.37 | 3.345 (2.251–4.973) | Adjusted; postoperative selection and operative covariates |
| Uguz et al. 2022 [13] | Postoperative AF | Continuous OR; apparent raw-unit scale | 1.007 (1.002–1.012) | Adjusted; small event count and conflicting event-count/specificity reporting |
| Önder et al. 2026 [31] | In-hospital death | OR for preoperative log-SII | 1.261 (1.052–1.512) | Includes postoperative length-of-stay and day-7 variables; log base and mortality event count unclear |
| Öntaş and Gözüaçık Rüzgar 2026 [20] | Wound infection | OR per 100 SII units | 0.93 (0.86–1.00) | Adjusted model mapping unclear; inverse direction retained |
| Vezir and Ertürk Tekin 2026 [37] | Pulmonary composite | OR per 100 SII units | 1.010 (1.006–1.014) | Source explicitly reports a 100-unit increment; postoperative selection and collinearity concerns |
| Dey et al. 2021 [10] | Composite poor outcome | Continuous OR; increment not explicit | 1.010 (1.003–1.016) | Adjusted; not a high-versus-low contrast |
| Parmana et al. 2024 [16] | Composite prolonged recovery | OR; ≥528.715 vs <528.715 | 3.645 (2.801–4.743) | Adjusted for operative duration/CPB; different composite |
| Sanrı and Aslan 2026 [32] | Hospital stay >7 days | OR | 1.001 (0.999–1.003) | No clear adjusted association; P=0.521; exposure coding insufficiently specified |
| Abanoz and Engin 2021 [26] | Early MACCE | Continuous SII OR | 3.88 (2.69–6.15) | Continuous coding confirmed; narrative synthesis only |
| Atasoy and Guven 2025 [41] | Postoperative AF | Reported multivariable SII OR; scaling unclear | 2.041 (0.656–6.354) | P=0.218; correlated blood counts/indices considered in modeling; narrative only |
| Şahin and Yeniay 2026 [44] | In-hospital mortality | OR per SD increase in log-SII | 1.02 (0.79–1.31) | Adjusted for EuroSCORE II and preoperative base excess; retained descriptively and not included in the HR synthesis |
| Şahin and Yeniay 2026 [44] | One-year mortality | HR per SD increase in log-SII | 1.05 (0.86–1.29) | Adjusted for EuroSCORE II and preoperative base excess; included in the exploratory continuous mortality HR synthesis |
| Yüksel et al. 2024 [45] | Postoperative AF | Reported multivariable SII OR; increment/scaling unclear | 1.0502 (0.9735–1.1305) | P=0.11; age and mathematically related inflammatory variables included; narrative synthesis only |

These are selected source-reported individual associations, not pooled independent prognostic effects. Rounded or incompletely specified estimates are retained descriptively; some additional reported models could not be quantified reliably. OR, odds ratio; HR, hazard ratio; SD, standard deviation; CPB, cardiopulmonary bypass.

**Table 3.** Pooled associations between preoperative SII and clinical outcomes.

| Outcome | Synthesis status | k | n | Effect estimate (95% CI) | I<sup>2</sup> |
| --- | --- | --- | --- | --- | --- |
| Postoperative atrial fibrillation | Crude categorical association; two off-pump cohorts | 2 | 1,378 | 1.60 (0.53–4.82) | 68% |
| Perioperative IABP use | Exploratory support endpoint; crude | 2 | 1,378 | 2.45 (1.16–5.18) | 51% |
| Postoperative atrial fibrillation | Exploratory; includes unspecified adjustment | 3 | 2,434 | 1.90 (0.68–5.30) | 42.6% |
| Perioperative IABP use | Exploratory; includes unspecified adjustment | 3 | 2,434 | 3.62 (0.47–27.81) | 79.0% |
| Perioperative stroke | Exploratory; includes unspecified adjustment | 2 | 1,427 | 1.46 (0.57–3.73) | 0.0% |
| Perioperative myocardial infarction | Exploratory; includes unspecified adjustment | 2 | 1,427 | 2.57 (0.62–10.73) | 40.6% |
| All-cause mortality over differing follow-up horizons | Exploratory adjusted continuous association; per study-specific SD of log-SII | 2 | 9,611* | HR 1.11 (1.05–1.18) | 0.0% |

Şahin and Yeniay [44] evaluated 618 patients undergoing isolated CABG. Each SD increase in log-transformed preoperative SII was not independently associated with in-hospital mortality after adjustment for EuroSCORE II and preoperative base excess (OR 1.02, 95% CI 0.79–1.31; P=0.889). The corresponding one-year mortality HR was 1.05 (95% CI 0.86–1.29; P=0.604). ICU mortality was identical to in-hospital mortality in this cohort because no additional deaths occurred between ICU and hospital discharge. In-hospital and 30-day mortality were considered separately from longer-term mortality. In the off-pump Parmana cohort, 30-day deaths occurred in 3/63 patients with SII ≥878.057 and 12/308 patients with lower SII, yielding a calculated crude OR of 1.23 (95% CI 0.34–4.50) [15]. The on-pump Parmana report provided an OR of 3.505 (95% CI 1.883–6.523) for SII ≥528.715, without clearly specifying endpoint-specific covariate adjustment [16].

For in-hospital mortality, Önder et al. [31] reported an adjusted OR of 1.261 (95% CI 1.052–1.512; P=0.015) for log-transformed preoperative SII. The model also included postoperative length-of-stay and day-7 laboratory variables; the logarithmic transformation was insufficiently specified, and mortality event counts were not clearly reported. Toprak et al. [33] reported a strongly rounded baseline OR from a mortality-enriched sample of 51 deaths and 194 sampled survivors. The 51/245 proportion was therefore not treated as mortality incidence; the source cohort contained 874 eligible patients. For extended follow-up, Jacquemyn et al. reported an adjusted HR of 1.12 (95% CI 1.05–1.19) per standard-deviation increase in log-SII over a median 5.3 years [19]. In the diabetic Polish subgroup, preoperative SII >665 yielded an imprecise unadjusted mortality estimate (HR 2.45, 95% CI 0.77–7.85; P=0.131). The significant adjusted SII association reported in that study concerned postoperative measurements and was excluded from the preoperative synthesis [34]. The other Polish reports found no statistically significant difference in preoperative SII between subsequent survivors and nonsurvivors (P=0.106 and P=0.174) [35, 36] (Table 2; Supplementary Table S5).

An exploratory synthesis of the adjusted mortality HRs reported by Jacquemyn et al. and Şahin and Yeniay [18, 43] yielded a pooled HR of 1.11 (95% CI 1.05–1.18) per study-specific SD increase in log-transformed preoperative SII, using REML with a Wald CI (I^2^=0%; τ^2^=0; Figure 2C). Follow-up was one year in Şahin and Yeniay and a median of 5.3 years in Jacquemyn et al.; the latter contributed 91.3% of the meta-analytic weight. A modified Hartung–Knapp sensitivity analysis yielded a wider interval that included the null (HR 1.11, 95% CI 0.76–1.64; Supplementary Table S7). The pooled estimate therefore requires cautious interpretation despite the estimated I^2^ of 0%.

### 3.4 Postoperative atrial fibrillation

Fifteen reports provided eligible preoperative SII evidence relevant to postoperative atrial fibrillation, although definitions and observation periods differed. Two off-pump cohorts supplied exact categorical counts at the same SII threshold of 878.057 [10, 15]. These included 1,378 patients and 180 atrial-fibrillation events. The pooled crude OR was 1.60 (95% CI 0.53–4.82; I^2^=68%; τ^2^=0.4653; Figure 2A; Table 3).

Including the on-pump Parmana report, which used a different SII threshold and did not clearly specify endpoint-specific covariate adjustment, yielded an exploratory three-report synthesis involving 2,434 patients [16]. The pooled OR was 1.90 (95% CI 0.68–5.30; I^2^=42.6%; Table 3; Supplementary Table S7).

Individual adjusted associations varied in their magnitude and reporting completeness. Uguz et al. reported a continuous SII OR of 1.007 (95% CI 1.002–1.012), although event-count and specificity reporting were inconsistent [13]. Selcuk et al. reported an OR of 1.002 (95% CI 1.001–1.002), but the model included postoperative ICU stay and the estimate was strongly rounded [11]. Ata and Abanoz reported an OR of 1.548 (95% CI 1.265–2.896), with insufficiently specified exposure scaling and uncertainty regarding the reported confidence interval [28]. Yilmaz et al. also reported a positive adjusted association [38]. Topal et al. described SII as an independent predictor without reporting its numerical adjusted estimate and confidence interval, whereas Erol and Civelek provided unadjusted group comparisons only [12, 29] (Table 2; Supplementary Table S5).

Four additional reports evaluated preoperative SII within broader biomarker analyses. Magoon et al. observed 151 AF events among 899 patients; the unadjusted association was positive, but the printed adjusted estimate was internally inconsistent and was excluded from quantitative synthesis [39]. Yüksel et al. observed 108 AF events among 427 patients. Preoperative SII was higher among patients developing AF, but the reported multivariable association was nonsignificant (OR 1.0502, 95% CI 0.9735–1.1305; P=0.11), with insufficiently specified exposure scaling [45]. Atasoy and Guven observed 31 AF events among 223 patients aged >18 to ≤50 years; their reported multivariable SII association was also nonsignificant, with unclear exposure scaling [41]. Osmanaj et al. reported an unadjusted OR of 1.07 per 100-unit increase in SII (95% CI 1.05–1.09) among 3,481 patients [42]. These findings were summarized descriptively and were not incorporated into the categorical meta-analyses (Table 2; Supplementary Tables S4 and S5).

### 3.5 Acute kidney injury

Two reports specifically evaluated postoperative acute kidney injury. Aykut et al. identified 160 cases among 624 patients using a seven-day creatinine-based KDIGO assessment. SII >651.37 was associated with AKI in the reported adjusted model (OR 3.35, 95% CI 2.25–4.97), which included preoperative renal and clinical variables as well as intraoperative covariates [14]. Göç and Bağış observed 11 cases among 50 patients using a 48-hour creatinine assessment; median SII was 1,186.3 in patients with AKI and 695.8 in those without AKI (P=0.006), but no adjusted SII estimate was reported [30]. A pooled prognostic effect was therefore not estimable. Four renal-replacement events were reported by Aykut et al., without a separate SII association [14] (Table 2; Supplementary Table S5).

### 3.6 Cardiovascular and cerebrovascular outcomes

Jacquemyn et al. reported an adjusted HR of 1.08 (95% CI 1.03–1.13) per standard-deviation increase in log-SII for long-term five-component MACCE [19]. Wang et al. included SII in a MACE model developed in 176 patients, but its HR and confidence interval were too strongly rounded for reliable variance estimation; performance of the full nomogram was not assigned to SII alone [17]. Abanoz and Engin [26] reported 34 early MACCE events among 297 patients and an adjusted continuous SII OR of 3.88 (95% CI 2.69–6.15). Continuous coding was confirmed, but predictor scaling and the reported confidence interval required clarification; the estimate was therefore retained descriptively. The different composites, horizons and SII scales precluded pooling these effects.

Dolapoglu and Avci evaluated 527 patients, of whom 103 developed MACCE during a reported median follow-up of 4.6 years. Preoperative SII was higher among patients with MACCE than among those without events (median 859 versus 627; P<0.001), and SII-alone discrimination was modest (AUC 0.668). The adjusted PIV association was not treated as an SII estimate [40].

Exploratory syntheses of perioperative stroke and myocardial infarction combined crude event counts from the off-pump Parmana cohort with adjustment-unspecified estimates from the on-pump report [15, 16]. Each synthesis included 1,427 patients. Pooled estimates were imprecise for stroke (OR 1.46, 95% CI 0.57–3.73; I^2^=0.0%) and myocardial infarction (OR 2.57, 95% CI 0.62–10.73; I^2^=40.6%) (Table 3; Supplementary Table S7).

Xue et al. evaluated 6,549 patients undergoing elective OPCABG across eight centers, with 136 postoperative cerebral infarctions. The reported SII comparator AUC was 0.5165 (95% CI 0.4658– 0.5672). Section 3.4 labeled the comparator “SII⁻¹,” whereas the discussion referred to SII; this ambiguity was retained in the extraction. Regression estimates for the principal systemic glucose–inflammation index were not assigned to SII [43].

### 3.7 Infection, pulmonary complications and delirium

The diabetic CABG wound-infection study reported 21 infections among 300 patients and an inverse association with preoperative SII: an unadjusted OR of 0.91 (95% CI 0.84–0.99) and a reported adjusted OR of 0.93 (95% CI 0.86–1.00), each per 100-unit increase in SII. The inverse direction was retained, although the adjusted covariate set and follow-up origin were inconsistently described [20]. Dey’s “other infection” endpoint was not considered equivalent to wound infection, and these findings were not pooled [10] (Supplementary Table S3).

Vezir and Ertürk Tekin reported 114 pulmonary-complication events among 1,034 patients and an adjusted OR of 1.010 (95% CI 1.006–1.014) per 100 SII units, as explicitly stated in the source. Selection requiring at least 48 hours of postoperative stay, correlated operative covariates and incompletely enumerated component criteria were relevant limitations [37]. Zhao et al. evaluated delirium in patients with prior cerebral infarction, observing 113 cases among 321 patients; both full-cohort and matched adjusted estimates had collapsed rounded confidence intervals, precluding reliable meta-analysis [18]. Each endpoint lacked a second compatible estimate (Table 2; Supplementary Table S5).

### 3.8 Postoperative recovery and support requirements

Reported associations with ventilation duration and ICU stay varied. Dey et al. described Pearson correlations of 0.676 and 0.527, respectively, while the hospital-stay correlation was 0.044 (P=0.165) [10]. Parmana’s off-pump cohort showed little correlation between SII and ventilation duration (r=0.044; P=0.402) or ICU stay (r=0.067; P=0.200), and a weak correlation with hospital stay (r=0.133; P=0.010) [15]. Correlation methods and complete-pair denominators were not consistently specified across reports [15, 30, 31] (Supplementary Table S5, Panel B). Parmana’s off-pump report provided bivariate RRs for prolonged ventilation and ICU stay, whereas the on-pump report provided ORs with unspecified endpoint-specific adjustment [15, 16]. These measures were not combined. Sanrı and Aslan reported a null adjusted SII association with hospital stay >7 days (OR 1.001, 95% CI 0.999–1.003; P=0.521), despite an unadjusted group difference; the regression coding remained unclear [32].

An exploratory synthesis of crude IABP-use counts from the Dey and off-pump Parmana cohorts included 1,378 patients and indicated greater IABP use among patients with higher preoperative SII (OR 2.45, 95% CI 1.16–5.18; I^2^=51%) [10, 15]. Adding the adjustment-unspecified estimate from the on-pump Parmana report increased the pooled OR to 3.62 but substantially widened its confidence interval (95% CI 0.47– 27.81; I^2^=79.0%) [16] (Table 3; Supplementary Table S7). Composite postoperative outcomes also differed. Dey et al. reported an adjusted continuous SII OR of 1.010 (95% CI 1.003–1.016) for a composite poor outcome, while Parmana’s on-pump report described an adjusted high-versus-low OR of 3.645 (95% CI 2.801–4.743) for a composite of mechanical ventilation >24 hours, ICU stay >48 hours or hospital stay >7 days [10, 16]. Alagha et al. reported an unadjusted association with a postoperative complication composite, but SII was not retained in the final adjusted model [27] (Table 2).

### 3.9 Discrimination and threshold reporting

Apparent SII discrimination for postoperative AF varied substantially, with reported AUCs ranging from 0.587 to 0.946 across seven reports [11–13, 28, 38, 39, 42]. Reported numerical thresholds ranged from 706.7 to 1066.23, although units and threshold derivation require consideration. Additional comparator analyses showed modest MACCE discrimination (AUC 0.668) and near-chance discrimination for in-hospital mortality (AUC 0.517), one-year mortality (AUC 0.516) and postoperative cerebral infarction (reported comparator AUC 0.5165) [40, 43, 44]. Osmanaj et al. reported small improvements when SII was added to a model comprising age, hypertension and left atrial diameter: NRI 0.04 (95% CI 0.01–0.06) and IDI 0.007 (95% CI 0.003–0.011) [42]. Discrimination measures were tabulated without pooling or reconstruction of exact classification counts from rounded percentages (Supplementary Table S4).

### 3.10 Methodological quality and risk of bias

NOS scores ranged from 5 to 9 across the 31 included reports, with a median of 7. Twenty-three reports scored at least 7, and eight scored between 5 and 6 (Supplementary Table S6). These totals were not interpreted as categorical risk-of-bias judgments. Important limitations included inadequate SII-specific confounding control, selected populations, incomplete outcome-availability reporting, correlated biomarker predictors and inconsistent statistical reporting.

### 3.11 Heterogeneity, sensitivity analyses and missing results

All pooled analyses contained only two or three reports. For atrial fibrillation, the common-effect estimate was 2.24 (95% CI 1.59–3.17), compared with the more uncertain random-effects estimate of 1.60 (95% CI 0.53–4.82). The crude-RR sensitivity analysis likewise remained imprecise (RR 1.51, 95% CI 0.61– 3.73). In the expanded three-report atrial-fibrillation analysis, leave-one-out estimates ranged from 1.42 to 2.10, and inference changed with the confidence-interval method and report omitted. The IABP association retained an interval above 1 when expressed as a crude RR (2.27, 95% CI 1.08–4.77), but became highly uncertain in the expanded synthesis. Full sensitivity results are provided in Supplementary Table S7.

## 4. Discussion

This review of 31 reports identified outcome-dependent associations between preoperative SII and clinical outcomes after CABG. Several studies reported adjusted associations, including the largest cohort’s associations with long-term mortality and MACCE. However, the updated evidence also included null adjusted mortality findings and limited discrimination for mortality and postoperative cerebral infarction. The count-based AF synthesis remained imprecise, while the crude IABP association remained exploratory. These findings support further evaluation of SII but do not establish a consistent independent prognostic effect, a clinically transferable threshold or reproducible benefit beyond established risk assessment.

Mortality associations depended on the time horizon and analytical approach. Jacquemyn et al. reported an adjusted hazard ratio of 1.12 (95% CI 1.05–1.19) per standard-deviation increase in log-SII for long-term mortality [19]. Önder et al. also reported an association with in-hospital mortality, although adjustment included postoperative variables [31]. Conversely, Polish off-pump studies provided imprecise or nonsignificant baseline SII findings [34–36]. These differences may reflect case mix, competing determinants of early and late death, and predictor coding. Early mortality pooling combined crude and adjustment-unspecified estimates and should therefore remain exploratory; it cannot resolve whether SII contributes information beyond established baseline risk. Şahin and Yeniay did not provide clear evidence of an adjusted association with in-hospital or one-year mortality after accounting for EuroSCORE II and preoperative base excess [44]. Their one-year HR and Jacquemyn’s multiyear HR shared a standardized log-SII scale but differed in follow-up and covariate adjustment. The exploratory pooled adjusted HR suggested a positive association, This finding should be interpreted as an uncertain summary association across differing follow-up horizons.

Postoperative atrial fibrillation was the most frequently examined complication. Positive adjusted associations in several smaller cohorts contrasted with substantial heterogeneity and imprecision in our count-based synthesis [11, 13, 38]. Yüksel et al. reported higher SII among patients developing AF but no statistically significant multivariable association; interpretation was complicated by the inclusion of related inflammatory variables, including MII-3, which incorporates SII directly [45]. In the largest cohort, the adjusted multimarker analysis was nonsignificant for atrial fibrillation; this was not an isolated SII-specific estimate [19]. Differences in monitoring, observation periods, ventricular function, and covariate selection may contribute to the inconsistency [46]. The pooled CI does not demonstrate absence of an association, but it limits confidence in its magnitude. Moreover, apparently high discrimination in selected derivation cohorts should not be equated with reproducible individual risk prediction.

The renal findings were directionally concordant: Aykut et al. reported an adjusted association between high SII and acute kidney injury, and Göç and Bağış observed higher baseline SII among affected patients [14, 30]. Different ascertainment windows and the absence of a second compatible adjusted estimate prevented pooling. Pulmonary complications and delirium also showed individual-study signals, with restricted populations or reporting limitations [18, 37]. The inverse wound-infection association in diabetic patients and the null adjusted hospital-stay association challenge a uniform relationship across outcomes [20, 32]. The infection findings warrant verification and replication rather than interpretation as a protective biological effect.

SII combines platelet, neutrophil and lymphocyte counts and may reflect aspects of the baseline inflammatory and immune state [7]. Cardiopulmonary bypass and ischemia–reperfusion can provoke immune activation and organ injury [6, 47]. Higher preoperative SII may therefore identify patients with greater susceptibility to surgical stress, although this remains a mechanistic interpretation. Cell counts do not directly measure cellular activation or immune competence, and similar SII values can arise from different cellular profiles. Preoperative measurements retain relevance for baseline assessment, whereas later measurements may reflect operative exposure, treatment or evolving complications.

SII is readily calculated from routine blood counts, making it a practical candidate for evaluation alongside established clinical risk models. Nevertheless, inexpensive measurement does not establish clinical usefulness. Osmanaj et al. reported small improvements beyond a limited AF model comprising age, hypertension and left atrial diameter, but this does not establish added value beyond comprehensive surgical risk models or external reproducibility [42]. Şahin and Yeniay found little prognostic contribution from SII for mortality, while Xue et al. reported limited stroke discrimination [43, 44]. Current evidence therefore does not justify selecting an operative technique, prescribing prophylaxis or delaying CABG on the basis of an SII cutoff alone. Future evaluations should report calibration, discrimination and clinical net benefit, followed by external validation.

Strengths include restriction to preoperative exposure, outcome-specific synthesis, separation of early from long-term endpoints, and explicit distinction between crude and adjusted associations. Incompatible predictor scales, effect measures, and composites were not combined. Rounded confidence intervals were not converted into spurious precision, and composite-model performance was not attributed to SII alone. These decisions preserve interpretability despite reducing the number of pooled studies.

The evidence remains observational and predominantly retrospective. Residual confounding, small event counts, and adjustment for downstream variables affect interpretation. Sampling intervals, thresholds, outcome definitions, and follow-up differed; same-sample cutoff selection may overestimate performance. Possible cohort overlap means that the aggregate sample cannot be interpreted as unique participants. These limitations reduce confidence in the magnitude and generalizability of the reported associations.

Prospective multicenter cohorts should prespecify sampling windows, standardize units and endpoints, and model SII continuously with assessment of nonlinearity. Analyses should adjust for clinically relevant baseline factors and distinguish preoperative prediction from models incorporating operative or postoperative information. Adequate event-based sample sizes, transparent missing-data handling, and optimism correction are essential [22, 48]. External validation should compare SII with NLR, PLR, SIRI, individual counts, and established clinical models, reporting calibration, discrimination, and net benefit. If reproducible incremental value is demonstrated, subsequent impact studies should test whether SII-informed care improves outcomes. Individual-participant-data collaboration could help harmonize scales and resolve overlap.

## 5. Conclusion

Preoperative SII was associated with selected outcomes after CABG, but findings and discriminatory performance varied across endpoints and follow-up horizons. The categorical AF synthesis remained imprecise, and the exploratory IABP association requires cautious interpretation. The exploratory adjusted mortality synthesis suggested a positive association, although inference was sensitive to the CI method and dominated by one cohort. Reproducible incremental value beyond established clinical risk models and clinically transferable thresholds remain unestablished. Further studies should standardize preoperative measurement, transparently model SII and evaluate its added value through external validation.

## Data Availability

The data supporting the findings of this systematic review and meta-analysis are available from the corresponding author upon reasonable request.

## Abbreviations

AF: Atrial fibrillation
AKI: Acute kidney injury
AUC: Area under the receiver operating characteristic curve
CABG: Coronary artery bypass grafting
CI: Confidence interval
CPB: Cardiopulmonary bypass
EF: Ejection fraction
EuroSCORE II: European System for Cardiac Operative Risk Evaluation II
HFpEF: Heart failure with preserved ejection fraction
HR: Hazard ratio
IABP: Intra-aortic balloon pump
ICU: Intensive care unit
IQR: Interquartile range
KDIGO: Kidney Disease: Improving Global Outcomes
MACE: Major adverse cardiovascular events
MACCE: Major adverse cardiovascular and cerebrovascular events
MI: Myocardial infarction
NLR: Neutrophil-to-lymphocyte ratio
NOS: Newcastle–Ottawa Scale
NR: Not reported
OPCAB: Off-pump coronary artery bypass
OR: Odds ratio
PLR: Platelet-to-lymphocyte ratio
PRISMA: Preferred Reporting Items for Systematic Reviews and Meta-Analyses
PRISMA-S: PRISMA extension for reporting literature searches in systematic reviews
REML: Restricted maximum likelihood
ROC: Receiver operating characteristic
RR: Risk ratio
SD: Standard deviation
SII: Systemic immune-inflammation index
SIRI: Systemic inflammation response index
STS: Society of Thoracic Surgeons

## Declarations

### Ethics approval and consent to participate

This systematic review and meta-analysis used aggregate data from published studies and did not involve direct recruitment of participants or access to identifiable individual-level data. Ethics approval and consent to participate were therefore not required.

### Consent for publication

Not applicable.

### Authors’ contributions

S.N. and K.J.R. conceptualized the study and developed the review protocol. M.M.M. and M.G. conducted the literature searches. M.G. and K.J.R. screened records and assessed study eligibility, with M.M.M. adjudicating disagreements. N.T. and M.H.N. extracted data, with S.N. adjudicating unresolved disagreements. M.G. and M.M.M. assessed methodological quality. S.N. and M.M.A. performed the statistical analyses. A.R.N., K.J.R., and M.G. drafted the manuscript. All authors contributed to interpretation of the findings and critical manuscript revision, approved the final manuscript, and agreed to be accountable for the work.

## Acknowledgements

Not applicable.

