## Supplementary Material for "Preoperative Systemic Immune-Inflammation Index and Clinical Outcomes After Coronary Artery Bypass Grafting: A Systematic Review and Meta-analysis"

**Supplementary Table S1 PRISMA 2020 checklist**

| **Section and Topic** | **Item #** | **Checklist item** | **Location where item is reported** |
| --- | --- | --- | --- |
| **TITLE** | | |  |
| Title | 1 | Identify the report as a systematic review. | p 1 |
| **ABSTRACT** | | |  |
| Abstract | 2 | See the PRISMA 2020 for Abstracts checklist. | p 2 |
| **INTRODUCTION** | | |  |
| Rationale | 3 | Describe the rationale for the review in the context of existing knowledge. | pp 3–4 |
| Objectives | 4 | Provide an explicit statement of the objective(s) or question(s) the review addresses. | pp 3–4 |
| **METHODS** | | |  |
| Eligibility criteria | 5 | Specify the inclusion and exclusion criteria for the review and how studies were grouped for the syntheses. | pp 4–5, 7–8 |
| Information sources | 6 | Specify all databases, registers, websites, organisations, reference lists and other sources searched or consulted to identify studies. Specify the date when each source was last searched or consulted. | pp 5–6 |
| Search strategy | 7 | Present the full search strategies for all databases, registers and websites, including any filters and limits used. | pp 5–6; Supplementary pp 3–4 |
| Selection process | 8 | Specify the methods used to decide whether a study met the inclusion criteria of the review, including how many reviewers screened each record and each report retrieved, whether they worked independently, and if applicable, details of automation tools used in the process. | p 6 |
| Data collection process | 9 | Specify the methods used to collect data from reports, including how many reviewers collected data from each report, whether they worked independently, any processes for obtaining or confirming data from study investigators, and if applicable, details of automation tools used in the process. | pp 6–7 |
| Data items | 10a | List and define all outcomes for which data were sought. Specify whether all results that were compatible with each outcome domain in each study were sought (e.g. for all measures, time points, analyses), and if not, the methods used to decide which results to collect. | pp 5–7 |
|  | 10b | List and define all other variables for which data were sought (e.g. participant and intervention characteristics, funding sources). Describe any assumptions made about any missing or unclear information. | pp 5–7 |
| Study risk of bias assessment | 11 | Specify the methods used to assess risk of bias in the included studies, including details of the tool(s) used, how many reviewers assessed each study and whether they worked independently, and if applicable, details of automation tools used in the process. | p 7 |
| Effect measures | 12 | Specify for each outcome the effect measure(s) (e.g. risk ratio, mean difference) used in the synthesis or presentation of results. | pp 7–8 |
| Synthesis methods | 13a | Describe the processes used to decide which studies were eligible for each synthesis (e.g. tabulating the study intervention characteristics and comparing against the planned groups for each synthesis (item #5)). | pp 7–8 |
|  | 13b | Describe any methods required to prepare the data for presentation or synthesis, such as handling of missing summary statistics, or data conversions. | p 7 |
|  | 13c | Describe any methods used to tabulate or visually display results of individual studies and syntheses. | p 8 |
|  | 13d | Describe any methods used to synthesize results and provide a rationale for the choice(s). If meta-analysis was performed, describe the model(s), method(s) to identify the presence and extent of statistical heterogeneity, and software package(s) used. | pp 7–8 |
|  | 13e | Describe any methods used to explore possible causes of heterogeneity among study results (e.g. subgroup analysis, meta-regression). | Not reported |
|  | 13f | Describe any sensitivity analyses conducted to assess robustness of the synthesized results. | p 8 |
| Reporting bias assessment | 14 | Describe any methods used to assess risk of bias due to missing results in a synthesis (arising from reporting biases). | Not reported |
| Certainty assessment | 15 | Describe any methods used to assess certainty (or confidence) in the body of evidence for an outcome. | Not reported |
| **RESULTS** | | |  |
| Study selection | 16a | Describe the results of the search and selection process, from the number of records identified in the search to the number of studies included in the review, ideally using a flow diagram. | pp 8, 24 |
|  | 16b | Cite studies that might appear to meet the inclusion criteria, but which were excluded, and explain why they were excluded. | Not reported |
| Study characteristics | 17 | Cite each included study and present its characteristics. | pp 8–9, 20–21 |
| Risk of bias in studies | 18 | Present assessments of risk of bias for each included study. | p 12; Supplementary pp 10–11 |
| Results of individual studies | 19 | For all outcomes, present, for each study: (a) summary statistics for each group (where appropriate) and (b) an effect estimate and its precision (e.g. confidence/credible interval), ideally using structured tables or plots. | pp 9–12, 22; Supplementary pp 4–9 |
| Results of syntheses | 20a | For each synthesis, briefly summarise the characteristics and risk of bias among contributing studies. | pp 8–12, 20–23; Supplementary pp 10–11 |
|  | 20b | Present results of all statistical syntheses conducted. If meta-analysis was done, present for each the summary estimate and its precision (e.g. confidence/credible interval) and measures of statistical heterogeneity. If comparing groups, describe the direction of the effect. | pp 9–12, 23, 25–26; Supplementary pp 12–14 |
|  | 20c | Present results of all investigations of possible causes of heterogeneity among study results. | Not reported |
|  | 20d | Present results of all sensitivity analyses conducted to assess the robustness of the synthesized results. | p 13; Supplementary pp 12–14 |
| Reporting biases | 21 | Present assessments of risk of bias due to missing results (arising from reporting biases) for each synthesis assessed. | Not reported |
| Certainty of evidence | 22 | Present assessments of certainty (or confidence) in the body of evidence for each outcome assessed. | Not reported |
| **DISCUSSION** | | |  |
| Discussion | 23a | Provide a general interpretation of the results in the context of other evidence. | pp 13–14 |
|  | 23b | Discuss any limitations of the evidence included in the review. | pp 13–15 |
|  | 23c | Discuss any limitations of the review processes used. | p 15 |
|  | 23d | Discuss implications of the results for practice, policy, and future research. | pp 14–15 |
| **OTHER INFORMATION** | | |  |
| Registration and protocol | 24a | Provide registration information for the review, including register name and registration number, or state that the review was not registered. | p 4 |
|  | 24b | Indicate where the review protocol can be accessed, or state that a protocol was not prepared. | p 4 |
|  | 24c | Describe and explain any amendments to information provided at registration or in the protocol. | Not reported |
| Support | 25 | Describe sources of financial or non-financial support for the review, and the role of the funders or sponsors in the review. | p 17 |
| Competing interests | 26 | Declare any competing interests of review authors. | p 17 |
| Availability of data, code and other materials | 27 | Report which of the following are publicly available and where they can be found: template data collection forms; data extracted from included studies; data used for all analyses; analytic code; any other materials used in the review. | p 17; Supplementary pp 4–14 |

**Supplementary Table S2 Database search strategies**

| Database | Search Strategy |
| --- | --- |
| PubMed | ( "systemic immune inflammation index"[tiab] OR "systemic immune inflammatory index"[tiab] OR "systemic immunoinflammation index"[tiab] OR "systemic immunoinflammatory index"[tiab] OR "systemic inflammation index"[tiab] OR SII[tiab] OR ( systemic[tiab] AND immun*[tiab] AND inflamm*[tiab] AND index[tiab] ) ) AND ( "Coronary Artery Bypass"[Mesh] OR "Coronary Artery Bypass, Off-Pump"[Mesh] OR CABG[tiab] OR OPCAB[tiab] OR OPCABG[tiab] OR "coronary bypass"[tiab] OR "coronary artery bypass"[tiab] OR "aortocoronary bypass"[tiab] OR "aorto-coronary bypass"[tiab] OR ( coronary[tiab] AND ( bypass*[tiab] OR surg*[tiab] OR revasculari*[tiab] ) ) ) |
| Embase | ( 'systemic immune inflammation index':ti,ab,kw OR 'systemic immune inflammatory index':ti,ab,kw OR 'systemic immunoinflammation index':ti,ab,kw OR 'systemic immunoinflammatory index':ti,ab,kw OR 'systemic inflammation index':ti,ab,kw OR sii:ti,ab,kw OR ( systemic:ti,ab,kw AND immun*:ti,ab,kw AND inflamm*:ti,ab,kw AND index:ti,ab,kw ) ) AND ( 'coronary artery bypass graft'/exp OR cabg:ti,ab,kw OR opcab:ti,ab,kw OR opcabg:ti,ab,kw OR 'coronary bypass':ti,ab,kw OR 'coronary artery bypass':ti,ab,kw OR 'aortocoronary bypass':ti,ab,kw OR 'aorto-coronary bypass':ti,ab,kw OR ( coronary:ti,ab,kw AND ( bypass*:ti,ab,kw OR surg*:ti,ab,kw OR revasculari*:ti,ab,kw ) ) ) |
| Scopus | TITLE-ABS-KEY ( "systemic immune inflammation index" OR "systemic immune inflammatory index" OR "systemic immunoinflammation index" OR "systemic immunoinflammatory index" OR "systemic inflammation index" OR SII OR ( systemic AND immun* AND inflamm* AND INDEX ) ) AND TITLE-ABS-KEY ( CABG OR OPCAB OR OPCABG OR "coronary bypass" OR "coronary artery bypass" OR "aortocoronary bypass" OR "aorto-coronary bypass" OR ( coronary AND ( bypass* OR surg* OR revasculari* ) ) ) |
| Web of Science | "systemic immune inflammation index" OR "systemic immune inflammatory index" OR "systemic immunoinflammation index" OR "systemic immunoinflammatory index" OR "systemic inflammation index" OR SII OR (systemic AND immun* AND inflamm* AND index) (All Fields) AND CABG OR OPCAB OR OPCABG OR "coronary bypass" OR "coronary artery bypass" OR "aortocoronary bypass" OR "aorto-coronary bypass" OR (coronary AND (bypass* OR surg* OR revasculari*)) (All Fields) |

**Supplementary Table S3. Individual inputs for quantitative syntheses**

**Panel A. Event counts and calculated crude odds ratios**

| Report | Outcome | Cutoff | High events/total | Low events/total | Crude OR (95% CI) |
| --- | --- | --- | --- | --- | --- |
| Dey et al. 2021 [1] | Postoperative AF | 878.057 | 62/239 | 96/768 | 2.45 (1.71–3.51) |
| Dey et al. 2021 [1] | High vasoactive-inotropic support | 878.057 | 98/239 | 82/768 | 5.81 (4.12–8.21) |
| Dey et al. 2021 [1] | IABP requirement | 878.057 | 20/239 | 20/768 | 3.42 (1.80–6.46) |
| Dey et al. 2021 [1] | Other infection | 878.057 | 50/239 | 10/768 | 20.05 (9.98–40.28) |
| Parmana et al. 2023 [2] | Postoperative AF | 878.057 | 3/63 | 19/308 | 0.76 (0.22–2.65) |
| Parmana et al. 2023 [2] | Stroke | 878.057 | 1/63 | 2/308 | 2.47 (0.22–27.64) |
| Parmana et al. 2023 [2] | Myocardial infarction | 878.057 | 2/63 | 8/308 | 1.23 (0.25–5.93) |
| Parmana et al. 2023 [2] | Mortality | 878.057 | 3/63 | 12/308 | 1.23 (0.34–4.50) |
| Parmana et al. 2023 [2] | Perioperative IABP use | 878.057 | 8/63 | 26/308 | 1.58 (0.68–3.67) |

All contrasts compare SII ≥878.057 with SII <878.057. Events/total are source-reported counts; ORs and 95% CIs are calculated from the corresponding 2×2 tables. AF and IABP rows from both reports contribute to the two-report crude syntheses. Parmana 2023 mortality, stroke and myocardial-infarction rows also contribute to the exploratory analyses with Parmana 2024. High vasoactive-inotropic support and other infection are descriptive contrasts and were not pooled. AF, atrial fibrillation; IABP, intra-aortic balloon pump; OR, odds ratio; CI, confidence interval.

**Panel B. Reported on-pump effects used in exploratory syntheses**

| Report | Outcome | SII comparison | Cohort n | Reported OR (95% CI) |
| --- | --- | --- | --- | --- |
| Parmana et al. 2024 [3] | **Postoperative atrial fibrillation** | **≥528.715 vs <528.715** | **1,056** | **1.721 (1.115–2.656)** |
| Parmana et al. 2024 [3] | **IABP use** | **≥528.715 vs <528.715** | **1,056** | **8.309 (4.072–16.957)** |
| Parmana et al. 2024 [3] | **30-day mortality** | **≥528.715 vs <528.715** | **1,056** | **3.505 (1.883–6.523)** |
| Parmana et al. 2024 [3] | **Stroke** | **≥528.715 vs <528.715** | **1,056** | **1.325 (0.477–3.682)** |
| Parmana et al. 2024 [3] | **Myocardial infarction** | **≥528.715 vs <528.715** | **1,056** | **5.277 (1.135–24.539)** |

**Panel C. Adjusted continuous mortality estimates used in the exploratory HR synthesis**

| Report | Outcome | Exposure scale | Adjusted HR (95% CI) | Follow-up |
| --- | --- | --- | --- | --- |
| Jacquemyn et al. 2026 [4] | All-cause mortality | Per study-specific SD of log-SII | 1.12 (1.05–1.19) | Median 5.3 years |
| Şahin and Yeniay 2026 [5] | All-cause mortality | Per study-specific SD of log-SII | 1.05 (0.86–1.29) | One year |

**Supplementary Table S4 Reported discrimination of preoperative SII alone**

| Report | Outcome | Cutoff | AUC (95% CI) | Sensitivity (%) | Specificity (%) |
| --- | --- | --- | --- | --- | --- |
| Dey et al. 2021 [1] | Composite poor outcome | 878.057 | 0.984 (0.978–0.99) | 97.6% | 91.0% |
| Selcuk et al. 2021 [6] | Postoperative AF | 807.8 | 0.7107 (CI NR) | 60.8% | 80.9% |
| Ata & Abanoz 2021 [7] | Postoperative AF | 986 | 0.784 (0.729–0.839) | 72.2% | 74.4% |
| Uguz et al. 2022 [8] | Postoperative AF | 866.04 | 0.946 (0.887–0.979) | 92.3% | Unclear* |
| Topal et al. 2022 [9] | Postoperative AF | 706.7 | 0.587 (0.54–0.634) | 86.6% | 29.3% |
| Yilmaz et al. 2023 [10] | Postoperative AF | 712.8 | 0.781 (0.727–0.835) | 85.0% | 61.2% |
| Aykut et al. 2023 [11] | Postoperative AKI | 651.37 | 0.718 (0.676–0.76) | 65.0% | 64.9% |
| Alagha et al. 2023 [12] | Complication composite | 603.08 | 0.632 (0.5–0.763) | 71.0% | 55.9% |
| Önder et al. 2026 [13] | In-hospital mortality | 610.3 | 0.631 (0.499–0.763) | 65.0% | 60.0% |
| Öntaş and Gözüaçık Rüzgar 2026 [14] | Wound infection | NR | 0.374 (0.312–0.436)† | NR | NR |
| Parmana et al. 2023 [2] | Prolonged ventilation | 600.897 | 0.658 (0.575–0.741) | 64.3% | 63.8% |
| Sanrı and Aslan 2026 [15] | Prolonged hospital stay | 653 | 0.646 (CI NR) | 75.0% | 59.0% |
| Parmana et al. 2024 [3] | Recovery composite | 528.715 | 0.669 (0.636–0.702) | 65.0% | 67.0% |
| Urbanowicz et al. 2022 [16] | Long-term mortality | 665 | 0.643 (CI NR) | 75.0% | 54.7% |
| Zhao et al. 2024 [17] | Postoperative delirium | NR | 0.73 (0.67–0.79) | NR | NR |
| Toprak et al. 2026 [18] | In-hospital mortality | 623.05 | 0.698 (0.622–0.774) | 86.3% | 49.0% |
| Vezir and Ertürk Tekin 2026 [19] | Pulmonary composite | 741.59 | 0.818 (0.77–0.867) | 64.0% | 87.4% |
| Abanoz and Engin 2021 [20] | Early MACCE | 912.4 | 0.864 (0.789–0.940) | 82.6% | 73.4% |
| Magoon et al. 2023 [21] | Postoperative AF | ≥1066.23 | 0.601; CI NR | 47.0 | 82.1 |
| Dolapoglu and Avci 2024 [22] | MACCE | NR | 0.668; CI NR | NR | NR |
| Osmanaj et al. 2026 [23] | Postoperative AF | NR in supplied main article | 0.61 (0.59–0.63) | 61.0 | 59.0 |
| Şahin and Yeniay 2026 [5] | In-hospital mortality | ≥721.9 | 0.517 (0.439–0.595) | 47.9 | 64.4 |
| Şahin and Yeniay 2026 [5] | One-year mortality | ≥840.6 | 0.516 (0.448–0.583) | 37.0 | 73.4 |
| Xue et al. 2026 [24] | Postoperative cerebral infarction | NR | 0.5165 (0.4658–0.5672) | NR | NR |

AUCs refer to SII alone for the stated endpoint, not to multivariable nomograms or individual components of composite outcomes. Source precision is retained. *Uguz et al. reported specificity of 1.1% in the text and 1.11% in Figure 1. These values are inconsistent with the accompanying sensitivity, Youden index and ROC curve. Specificity was therefore classified as unclear and was not used for quantitative synthesis. †The wound-infection AUC of 0.374 is retained in its reported direction, consistent with lower SII in the infection group; it has not been inverted. Cutoffs, scales and follow-up differ across reports. These values do not establish a universal threshold. AUC, area under the receiver operating characteristic curve; NR, not reported; AKI, acute kidney injury; MACCE, major adverse cardiovascular and cerebrovascular events.

**Supplementary Table S5 Unadjusted associations of preoperative SII with clinical outcomes**

Panel A SII distributions by clinical outcome

| Report | Outcome | Summary type | Outcome group | Comparison group | P |
| --- | --- | --- | --- | --- | --- |
| Dey et al. 2021 [1] | Composite poor outcome | Mean / SD | Poor outcome: 1262.11 ± 229.96 | Favorable outcome: 584.36 ± 189.44 | <0.001 |
| Selcuk et al. 2021[6] | Postoperative AF | Median / IQR | AF: 908 [455–1435] | No AF: 486 [353–716] | <0.001 |
| Ata & Abanoz 2021 [7] | Postoperative AF | Median / range | AF: 1034 (560–2496) | No AF: 878 (380–2134) | <0.001 |
| Uguz et al. 2022 [8] | Postoperative AF | Median / range | AF: 1144 (783–10236.1) | No AF: 562.1 (165.2–2305.4) | <0.001 |
| Topal et al. 2022 [9] | Postoperative AF | Median / range | AF: 1150.1 (199.2–5594.3) | No AF: 991.4 (175–4895.4) | 0.001 |
| Urbanowicz et al. 2022 [25] | All-cause mortality | Median / IQR | Non-survivor: 618 [498–1017] | Survivor: 562 [409–792] | 0.106 |
| Yilmaz et al. 2023 [10] | Postoperative AF (full cohort) | Median / IQR | AF: 1109 [720–2013] | No AF: 609 [373–754] | <0.001 |
| Yilmaz et al. 2023 [10] | Postoperative AF (matched) | Median / IQR | AF: 1109 [720–2013] | No AF: 628 [396–720] | <0.001 |
| Erol & Civelek 2025 [26] | Postoperative AF | Mean / SD | AF: 1188 ± 447 | No AF: 657 ± 1137 | 0.008 |
| Aykut et al. 2023 [11] | Postoperative AKI | Median / IQR | AKI: 747.18 [577.9–1043.2] | No AKI: 541.17 [388.5–758.8] | <0.001 |
| Alagha et al. 2023 [12] | Complication composite | Median / IQR | Poor outcome: 828.25 [461.65–1084.09] | Favorable outcome: 566.16 [478.39–748] | 0.036 |
| Önder et al. 2026 [13] | In-hospital mortality | Central value (interval); type NR* | Mortality (+): 83.7 (45.4–93.8) | Mortality (−): 53.4 (37.6–78.9) | 0.040 |
| Önder et al. 2026 [13] | Postoperative AF | Central value (interval); type NR* | AF (+): 94.6 (70.3–139.7) | AF (−): 47.9 (35.2–65.4) | <0.001 |
| Öntaş and Gözüaçık Rüzgar 2026 [14] | Wound infection | Mean / SD | Wound infection: 958.48 ± 493.49 | No wound infection: 1293.56 ± 758.15 | 0.047 |
| Sanrı and Aslan 2026 [15] | Prolonged hospital stay | Mean / SD | Hospital >7 days: 911 ± 410 | Hospital ≤7 days: 738 ± 306 | 0.015 |
| Urbanowicz et al. 2022 [16] | Long-term mortality | Median / IQR | Deaths: 839 [611–1068] | Survivors: 617 [422–904] | 0.059 |
| Zhao et al. 2024 [17] | Postoperative delirium | Median / IQR | Delirium: 386.8 [230.87–905.48] | No delirium: 318.65 [175.79–475.76] | 0.003 |
| Toprak et al. 2026 [18] | In-hospital mortality | Mean / SD | In-hospital death: 1010.5 ± 411.62 | Survived: 776.56 ± 594.64 | 0.001 |
| Urbanowicz et al. 2022 [27] | Long-term mortality | Median / IQR | Died during follow-up: 668 [445–982] | Survived: 618 [424–903] | 0.174 |
| Vezir and Ertürk Tekin 2026 [19] | Pulmonary composite | Mean / SD | PPC: 825 ± 248 | No PPC: 548 ± 164 | <0.001 |
| Göç and Bağış 2026 [28] | Acute kidney injury | Median / range | AKI: 1186.3 (792.5–1684.7) | No AKI: 695.8 (452.4–1045.6) | 0.006 |
| Abanoz and Engin 2021 [20] | Early MACCE | Median / range | MACCE, n=34: 1240 (630–3276) | No MACCE, n=263: 730 (540–2594) | <0.001 |
| Magoon et al. 2023 [21] | Postoperative AF | Median / IQR | AF, n=151: 1040 [874–1185] | No AF, n=748: 950 [900.95–1036.32] | <0.001 |
| Dolapoglu and Avci 2024 [22] | MACCE | Median / IQR | MACCE, n=103: 859 [585–1173] | No MACCE, n=424: 627 [489–830] | <0.001 |
| Atasoy and Guven 2025 [29] | Postoperative AF | Median / IQR | AF, n=31: 961 [615–2185] | No AF, n=192: 678 [443–943] | <0.001 |
| Şahin and Yeniay 2026 [5] | In-hospital mortality | Median / IQR | Death, n=73: 672.0 [412.3–959.0] | Survived, n=545: 619.0 [435.5–868.0] | 0.636 |
| Yüksel et al. 2024 [30] | Postoperative AF | Median / minimum–maximum | AF, n=108: 1204.7 (211.5–5120.1) | No AF, n=319: 987.6 (196.3–4895.4) | 0.001 |

Values are source-reported means ± SD, medians [IQR], or medians (range), as indicated. Matched and full-cohort Yilmaz analyses concern the same underlying cohort and are not independent evidence. *Önder’s values were retained as printed. SII scaling was inconsistently specified across tables, and the mean ±SD headings conflicted with the displayed interval format.

**Panel B Correlations with postoperative recovery and laboratory outcomes**

| Report | Outcome | Nominal n | Coefficient type | Reported coefficient | P |
| --- | --- | --- | --- | --- | --- |
| Dey et al. 2021 [1] | Ventilation duration | 1007 | Pearson | 0.676 | <0.001 |
| Dey et al. 2021 [1] | ICU length of stay | 1007 | Pearson | 0.527 | <0.001 |
| Dey et al. 2021 [1] | Hospital length of stay | 1007 | Pearson | 0.044 | 0.165 |
| Önder et al. 2026 [13] | Extubation time | NR | Pearson or Spearman; specific type NR | 0.479 | <0.001 |
| Önder et al. 2026 [13] | Hospital length of stay | NR | Pearson or Spearman; specific type NR | 0.47 | <0.001 |
| Önder et al. 2026 [13] | ICU length of stay | NR | Pearson or Spearman; specific type NR | 0.362 | <0.001 |
| Parmana et al. 2023 [2] | Mechanical ventilation duration, hours | 371 | Pearson or Spearman; specific type NR | 0.044 | 0.402 |
| Parmana et al. 2023 [2] | ICU length of stay, hours | 371 | Pearson or Spearman; specific type NR | 0.067 | 0.200 |
| Parmana et al. 2023 [2] | Hospital length of stay, days | 371 | Pearson or Spearman; specific type NR | 0.133 | 0.010 |
| Parmana et al. 2023 [2] | Vasoactive-inotropic score | 371 | Pearson or Spearman; specific type NR | 0.095 | 0.069 |
| Göç and Bağış 2026 [28] | Postoperative lactate | 50 | Pearson or Spearman; endpoint-specific type NR | 0.421 | 0.002 |
| Göç and Bağış 2026 [28] | ICU length of stay | 50 | Pearson or Spearman; endpoint-specific type NR | 0.368 | 0.009 |
| Göç and Bağış 2026 [28] | Ventilation duration | 50 | Pearson or Spearman; endpoint-specific type NR | 0.331 | 0.019 |
| Göç and Bağış 2026 [28] | Postoperative creatinine | 50 | Pearson or Spearman; endpoint-specific type NR | 0.394 | 0.005 |

Nominal n is the report-level cohort size where available; the number of complete pairs for each correlation was not consistently reported and should not be assumed equal to the cohort size. Önder’s endpoint-specific n was unavailable. Pearson/Spearman allocation is retained as unresolved where the source does not specify the coefficient for an endpoint. These unadjusted correlations were not pooled and do not establish independent prognostic value. ICU, intensive care unit.

**Panel C. Reported unadjusted regression associations**

| Report | Outcome | Effect and scale | Estimate (95% CI) | P |
| --- | --- | --- | --- | --- |
| Magoon et al. 2023 [21] | Postoperative AF | Ordinary-SII OR; reported raw-unit scale | 1.002 (1.001–1.003) | <0.001 |
| Osmanaj et al. 2026 [23] | Postoperative AF | OR per 100 SII units | 1.07 (1.05–1.09) | <0.001 |
| Şahin and Yeniay 2026 [5] | In-hospital mortality | OR per SD increase in log-SII | 1.06 (0.83–1.35) | 0.653 |
| Şahin and Yeniay 2026 [5] | One-year mortality | HR per SD increase in log-SII | 1.11 (0.91–1.35) | 0.298 |

**Supplementary Table S6 Newcastle–Ottawa Scale assessments**

| Report | Index outcome | Selection (0–4) | Comparability (0–2) | Outcome/exposure (0–3) | Total (0–9) |
| --- | --- | --- | --- | --- | --- |
| Dey et al. 2021 [1] | 30-day composite poor outcome | 4 | 2 | 3 | 9 |
| Selcuk et al. 2021 [6] | Postoperative AF | 4 | 0 | 2 | 6 |
| Ata & Abanoz 2021 [7] | In-hospital postoperative AF | 3 | 2 | 3 | 8 |
| Uguz et al. 2022 [8] | In-hospital postoperative AF | 4 | 2 | 2 | 8 |
| Topal et al. 2022 [9] | In-hospital postoperative AF | 4 | 1 | 3 | 8 |
| Urbanowicz et al. 2022 [25] | 8-year all-cause mortality | 4 | 0 | 3 | 7 |
| Yilmaz et al. 2023 [10] | In-hospital postoperative AF; full cohort | 4 | 1 | 3 | 8 |
| Erol & Civelek 2025 [26] | Postoperative AF during admission | 3 | 0 | 3 | 6 |
| Aykut et al. 2023 [11] | AKI within seven postoperative days | 3 | 2 | 3 | 8 |
| Alagha et al. 2023 [12] | 30-day composite poor postoperative outcome | 3 | 0 | 2 | 5 |
| Önder et al. 2026 [13] | In-hospital mortality | 4 | 1 | 2 | 7 |
| Öntaş and Gözüaçık Rüzgar 2026 [14] | Postoperative wound infection | 3 | 2 | 2 | 7 |
| Jacquemyn et al. 2026 [4] | Long-term all-cause mortality; secondary MACCE | 4 | 2 | 3 | 9 |
| Parmana et al. 2023 [2] | Prolonged ventilation / ICU stay | 4 | 0 | 3 | 7 |
| Sanrı and Aslan 2026 [15] | Hospital stay longer than seven days | 4 | 2 | 3 | 9 |
| Parmana et al. 2024 [3] | Composite prolonged MV/ICU/hospital stay | 4 | 1 | 3 | 8 |
| Urbanowicz et al. 2022 [16] | Long-term mortality in diabetic subgroup | 3 | 0 | 3 | 6 |
| Zhao et al. 2024 [17] | Delirium in the first five postoperative days | 3 | 2 | 3 | 8 |
| Toprak et al. 2026 [18] | Index-hospitalization mortality; baseline SII only | 3 | 0 | 2 | 5 |
| Wang et al. 2024 [31] | MACE; development cohort SII Cox estimate | 3 | 1 | 2 | 6 |
| Urbanowicz et al. 2022 [27] | Long-term all-cause mortality; baseline SII | 4 | 0 | 3 | 7 |
| Vezir and Ertürk Tekin 2026 [19] | Postoperative pulmonary complications during admission | 3 | 1 | 3 | 7 |
| Göç and Bağış 2026 [28] | Creatinine-defined AKI within 48 hours | 4 | 0 | 3 | 7 |
| Abanoz and Engin 2021 [20] | Early MACCE | 4 | 2 | 2 | 8 |
| Magoon et al. 2023 [21] | AF within seven postoperative days | 3 | 2 | 3 | 8 |
| Dolapoglu and Avci 2024 [22] | Follow-up MACCE | 4 | 0 | 3 | 7 |
| Atasoy and Guven 2025 [29] | AF through hospital discharge | 3 | 0 | 3 | 6 |
| Osmanaj et al. 2026 [23] | AF within seven postoperative days | 4 | 2 | 3 | 9 |
| Şahin and Yeniay 2026 [5] | In-hospital and one-year mortality | 4 | 2 | 3 | 9 |
| Xue et al. 2026 [24] | In-hospital postoperative cerebral infarction; SII comparator | 4 | 0 | 2 | 6 |
| Yüksel et al. 2024 [30] | Postoperative AF through discharge | 4 | 1 | 3 | 8 |

**Supplementary Table S7 Meta-analysis specifications and sensitivity analyses**

| Outcome and synthesis | Model or sensitivity analysis | k | Estimate (95% CI) | I² | CI method |
| --- | --- | --- | --- | --- | --- |
| AF — two-report crude synthesis | Primary REML; τ²=0.4653 | 2 | OR 1.60 (0.53–4.83) | 67.9% | Wald |
| AF — two-report crude synthesis | Common-effect model | 2 | OR 2.24 (1.59–3.17) | — | Wald |
| AF — two-report crude synthesis | Crude RR instead of OR (REML) | 2 | RR 1.51 (0.61–3.73) | 60.3% | Wald |
| IABP — two-report crude synthesis (exploratory) | Primary REML; τ²=0.1528 | 2 | OR 2.45 (1.16–5.18) | 51.2% | Wald |
| IABP — two-report crude synthesis (exploratory) | Common-effect model | 2 | OR 2.58 (1.55–4.29) | — | Wald |
| IABP — two-report crude synthesis (exploratory) | Crude RR instead of OR (REML) | 2 | RR 2.27 (1.08–4.77) | 58.5% | Wald |
| AF — expanded exploratory synthesis | Primary REML; τ²=0.0535 | 3 | OR 1.90 (0.68–5.30) | 42.6% | Hartung–Knapp |
| AF — expanded exploratory synthesis | Common-effect model | 3 | OR 2.02 (1.54–2.65) | — | Wald |
| AF — expanded exploratory synthesis | Paule–Mandel random-effects model | 3 | OR 1.81 (0.57–5.75) | 63.7% | Hartung–Knapp |
| AF — expanded exploratory synthesis | REML with alternative Wald CI | 3 | OR 1.90 (1.27–2.85) | 42.6% | Wald |
| AF — expanded exploratory synthesis | REML omitting Dey 2021 [1] | 2 | OR 1.42 (0.72–2.80) | 31.8% | Wald |
| AF — expanded exploratory synthesis | REML omitting Parmana 2023 [2] | 2 | OR 2.10 (1.49–2.96) | 34.0% | Wald |
| AF — expanded exploratory synthesis | REML omitting Parmana 2024 [3] | 2 | OR 1.60 (0.53–4.83) | 67.9% | Wald |
| 30-day mortality — exploratory synthesis | Common-effect model | 2 | OR 2.88 (1.65–5.05) | — | Wald |
| IABP — expanded exploratory synthesis | Primary REML; τ²=0.5174 | 3 | OR 3.62 (0.47–27.81) | 79.0% | Hartung–Knapp |
| IABP — expanded exploratory synthesis | Common-effect model | 3 | OR 3.83 (2.53–5.79) | — | Wald |
| IABP — expanded exploratory synthesis | Paule–Mandel random-effects model | 3 | OR 3.62 (0.47–27.82) | 79.5% | Hartung–Knapp |
| IABP — expanded exploratory synthesis | REML with alternative Wald CI | 3 | OR 3.62 (1.45–9.06) | 79.0% | Wald |
| IABP — expanded exploratory synthesis | REML omitting Dey 2021 [1] | 2 | OR 3.68 (0.72–18.73) | 88.5% | Wald |
| IABP — expanded exploratory synthesis | REML omitting Parmana 2023 [2] | 2 | OR 5.25 (2.20–12.54) | 69.8% | Wald |
| IABP — expanded exploratory synthesis | REML omitting Parmana 2024 [3] | 2 | OR 2.44 (1.15–5.17) | 51.2% | Wald |
| Stroke — exploratory synthesis | Primary REML; τ²=0.0000 | 2 | OR 1.46 (0.57–3.73) | 0.0% | Wald |
| Stroke — exploratory synthesis | Common-effect model | 2 | OR 1.46 (0.57–3.73) | — | Wald |
| Myocardial infarction — exploratory synthesis | Primary REML; τ²=0.4313 | 2 | OR 2.57 (0.62–10.73) | 40.6% | Wald |
| Myocardial infarction — exploratory synthesis | Common-effect model | 2 | OR 2.59 (0.86–7.78) | — | Wald |
| Adjusted continuous mortality | REML; τ²=0 | 2 | HR 1.11 (1.05–1.18) | 0.0% | Wald |
| Adjusted continuous mortality | Same REML estimate | 2 | HR 1.11 (0.76–1.64) | 0.0% | Modified Hartung–Knapp; variance floor 1, df=1 |

REML, restricted maximum likelihood; τ², estimated between-report variance on the log-effect scale; k, number of reports. Primary estimates match main Table 3. I² is derived from the random-effects variance estimate and typical within-report variance. Common-effect rows show — because no model-derived random-effects I² is reported; this does not imply absence of observed heterogeneity. For the five two-report categorical syntheses, Paule–Mandel estimates and CIs reproduced the primary results to the displayed precision; duplicate rows are omitted. The expanded AF and IABP models use Hartung–Knapp intervals in their primary analyses; alternative Wald intervals illustrate sensitivity to the interval methodExpanded categorical syntheses and categorical mortality, stroke and myocardial-infarction syntheses include an estimate from Parmana et al. (2024) with unspecified endpoint-specific adjustment and are exploratory. The separate adjusted continuous mortality synthesis combines study-reported adjusted HRs per study-specific SD increase in log-SII; follow-up horizons and covariate adjustment differ between reports.
